# Butyrate modifies epigenetic and immune pathways in peripheral mononuclear cells from children with neurodevelopmental disorders associated with chromatin dysregulation

**DOI:** 10.1101/2025.07.03.25330764

**Authors:** Jessica P Hayes, Velda X Han, Brooke A Keating, Hiroya Nishida, Erica Tsang, Xianzhong Lau, Ruwani Dissanayake, Melanie Wong, Carolyn Ellaway, Brian S Gloss, Shekeeb S Mohammad, Markus J Hofer, Peter Valtchev, Shrujna Patel, Russell C Dale

## Abstract

Pathogenic DNA variants in chromatin-related genes cause an important minority of neurodevelopmental disorders (NDDs). Epigenetic mechanisms, including chromatin regulation, are associated with NDD etiopathogenesis. Therapeutic strategies targeting chromatin dysregulation, such as histone deacetylase inhibition with butyrate, show promise; however, its effects remain poorly understood. We performed single-cell RNA sequencing (scRNA-seq) on 101,539 peripheral immune cells from four children with functionally impairing NDDs (median age 11.2 years,; 3 females): three with *de novo* pathogenic variants in chromatin-related genes (*KMT2D*, *CHD7*, and *MECP2*), and one without a monogenic diagnosis (non-monogenic), compared with two sex-matched healthy controls (median age 12.5 years, 1 female). Patient cells also underwent scRNA-seq after *in vitro* butyrate treatment. We identified 136 to 6909 differentially expressed genes (DEGs) across patient comparisons. Untreated patient cells showed dysregulation of ribosomal and immune pathways compared to controls. *KMT2D*, *CHD7*, and the non-monogenic patients exhibited downregulation of ribosomal pathways, and upregulation of immune pathways, whereas the *MECP2* patient displayed upregulation of ribosomal pathways and mixed regulation of immune pathways. Butyrate largely reversed these pathways, normalizing ribosomal and immune pathways in *KMT2D, CHD7,* and non-monogenic cells, with partial effects in *MECP2*. Overall, butyrate induced up-regulation of ribosome, GTPase, cytoskeletal, mitochondrial pathways, and down-regulation of epigenetic and immune pathways. This is the first study to demonstrate that butyrate modulates epigenetic and immune gene networks in monogenic and non-monogenic NDDs. The shared ribosomal-immune RNA signature suggests a common downstream mechanism of chromatin dysregulation, positioning butyrate as a promising therapeutic modulator across diverse NDDs.

## Introduction

*De novo* pathogenic DNA variants in chromatin-related genes cause an important minority of neurodevelopmental disorders (NDDs) [1]. However, NDDs in most children are not due to rare pathogenic DNA variants but are instead thought to arise from interactions between common genetic variations and environmental factors, associated with epigenetic and sometimes immune dysregulation [2]. Environmental exposures are proposed to leave epigenetic marks on the genome, influencing the expression of genes involved in neurodevelopment [3,4]. Epigenetic mechanisms, such as DNA methylation, histone modification, chromatin remodelling and non-coding RNAs, change gene expression without altering the DNA sequence [5]. DNA is wrapped around histone bodies to form chromatin, and histone modifications influence chromatin availability as euchromatin (open and available for gene transcription) or heterochromatin (closed and less available for gene transcription) [6]. Epigenetic changes due to genetic or environmental factors can disrupt tightly regulated gene networks essential for brain development and immune function, contributing to NDDs [7,8].

Increasing evidence points to a transcriptomic signature in patients with NDDs of dysregulated ribosomal and immune pathways, which may be the result of chromatin dysfunction [7,8]. We hypothesise that therapeutic strategies targeting epigenetic dysregulation may be a potential intervention for NDDs. Histone deacetylase (HDAC) enzymes remove acetyl groups from histones, resulting in chromatin condensation and transcriptional repression. Therefore, inhibiting these enzymes with HDAC inhibitors promotes an euchromatin structure, facilitating access for the transcriptional machinery and enhancing gene expression [9].

Butyrate is a short chain fatty acid derived from microbial fermentation of dietary fiber in the colon and has known HDAC inhibitory and anti-inflammatory properties [10]. We previously showed that the ketogenic diet, which increases the production of β-hydroxybutyrate, improved cognitive outcomes in a child with Kabuki syndrome (caused by *KMT2D* [a histone methyltransferase] mutation) [11]. Using single-cell RNA sequencing (scRNA-seq), we showed that ribosomal protein and immune pathways were dysregulated at baseline and normalised with a ketogenic diet in Kabuki syndrome. As both epigenetic and immune dysregulation are relevant to the pathophysiology of NDDs [12–16], butyrate is positioned as a promising therapeutic for NDDs.

Due to the ubiquitous role of chromatin in cell function, *de novo* pathogenic variations in chromatin-related genes cause multiorgan genetic conditions including NDDs [11,17,18], and patients can suffer infection-provoked neuroregression^19,20^. We investigated gene expression of peripheral immune cells from children with NDDs, including those with *de novo* DNA variation in key chromatin-related genes: *KMT2D* (Kabuki syndrome)*, CHD7* (CHARGE syndrome), *MECP2* (Rett syndrome), as well as one patient with severe non-monogenic NDD. These genes (*KMT2D, CHD7, MECP2*) are central to chromatin function and are ubiquitously expressed in most cells of the body, including the immune system and brain. We examined gene regulation using scRNA-seq of peripheral mononuclear cells in these patients at baseline, and after *in vitro* butyrate treatment. We hypothesised that *in vitro* butyrate treatment would modify gene expression and suppress immune dysregulation in patients with NDDs.

## Methods

### Participants

Four children (< 18 years of age) with functionally impairing NDDs were recruited to this study (median age 11.2 years, 3 females). Three female children had *de novo* pathogenic variants in the chromatin-related genes *KMT2D, CHD7,* and *MECP2,* the fourth male child did not have a definable monogenic variant using trio exome sequencing (non-monogenic). None of the patients had an infection in the two weeks before blood draw. Table 1 briefly outlines the genetic variant and clinical phenotype.

**Table 1.**
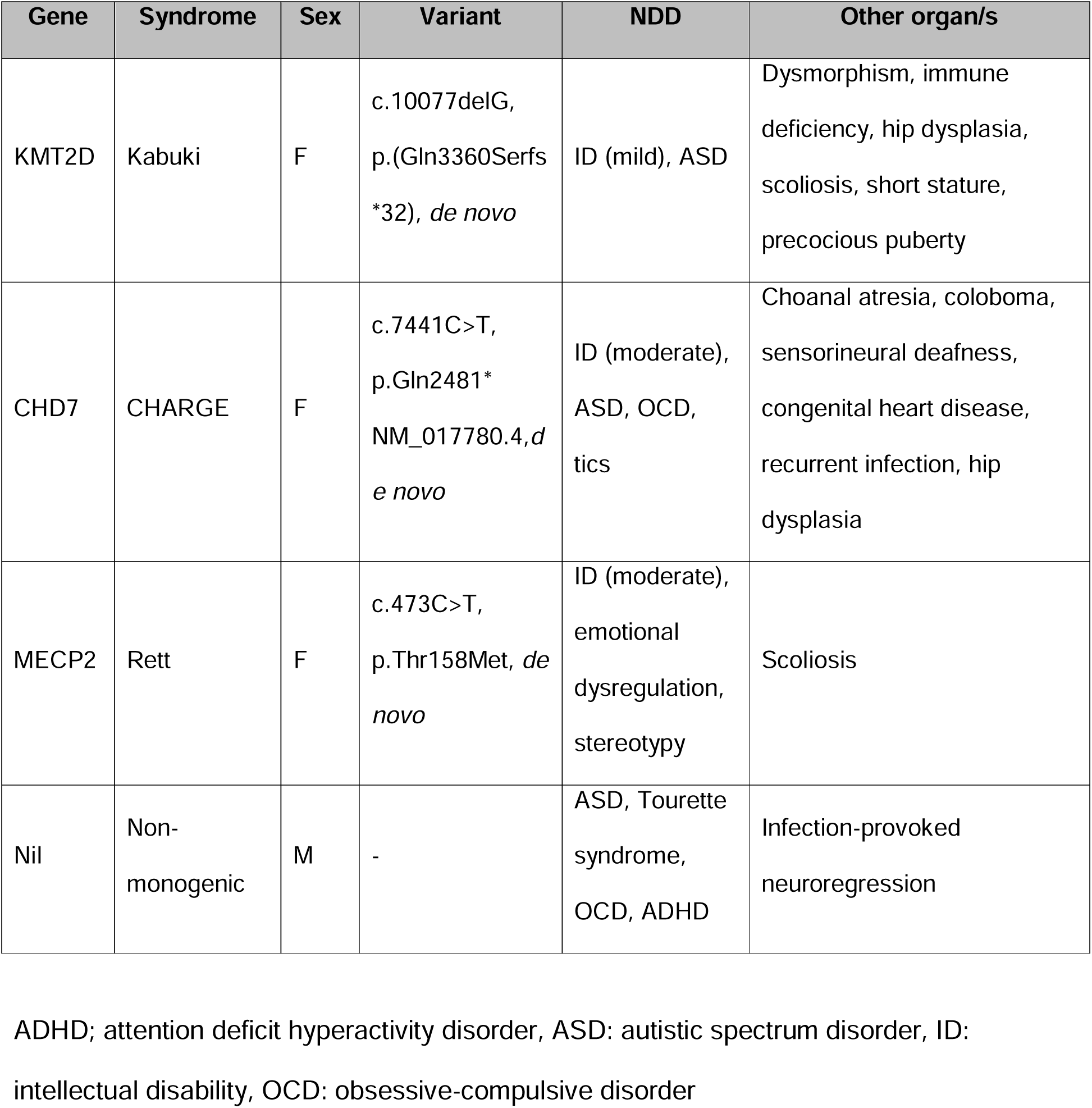
Patient syndrome, gene variant, and clinical phenotype.

| Gene | Syndrome | Sex | Variant | NDD | Other organ/s |
| --- | --- | --- | --- | --- | --- |
| KMT2D | Kabuki | F | c.10077delG,<br>p.(Gln3360Serfs<br>*32), *de novo* | ID (mild), ASD | Dysmorphism, immune<br>deficiency, hip dysplasia,<br>scoliosis, short stature,<br>precocious puberty |
| CHD7 | CHARGE | F | c.7441C>T,<br>p.Gln2481*<br>NM_017780.4,*d*<br>*e novo* | ID (moderate),<br>ASD, OCD,<br>tics | Choanal atresia, coloboma,<br>sensorineural deafness,<br>congenital heart disease,<br>recurrent infection, hip<br>dysplasia |
| MECP2 | Rett | F | c.473C>T,<br>p.Thr158Met, *de*<br>*novo* | ID (moderate),<br>emotional<br>dysregulation,<br>stereotypy | Scoliosis |
| Nil | Non-monogenic | M | - | ASD, Tourette<br>syndrome,<br>OCD, ADHD | Infection-provoked<br>neuroregression |

ADHD; attention deficit hyperactivity disorder, ASD: autistic spectrum disorder, ID: intellectual disability, OCD: obsessive-compulsive disorder

### Control selection

We recruited one age-matched healthy female for comparison with *KMT2D, CHD7* and *MECP2* patients, and one age-matched healthy male for comparison with the non-monogenic case. The controls were healthy children who had no neurodevelopmental or neuropsychiatric disorder, autoimmune diseases, severe allergic conditions, and no infection in the previous two weeks before blood draw from the child.

### Sample collection

Venous blood from the participants was collected in Vacutainer ACD tubes (BD Biosciences, BD367756) after gaining written consent. Peripheral blood mononuclear cells (PBMCs) were isolated and stored, as previously described [11].

### Treatment of PBMCs with butyrate

Frozen PBMC aliquots were thawed rapidly in a 37°C water bath, followed by washing with thawing medium consisting of RPMI 1640 supplemented with 10% foetal bovine serum (FBS), 1% GlutaMAX™, and 1% HEPES. The cells were then incubated in culture medium (RPMI 1640 supplemented with 10% FBS and 1% GlutaMAX™) at 37°C in a 5% CO_2_ atmosphere for 3 hours. Following the incubation, the cells were treated with 5mM sodium butyrate (Thermo Fisher Scientific) in culture media or control culture media (untreated) for 24 hours at 37°C in a 5% CO_2_ atmosphere. A dose of 5mM butyrate was chosen based on the ideal blood ketone ranges for the treatment of pediatric epilepsy with the ketogenic diet (2.0-5 mM β-hydroxybutyrate) [20] and butyrate has epigenetic effects at this concentration [11,21].

### HIVE^TM^ single-cell RNA sequencing

Following the 24-hour incubation with butyrate or control culture media, the cells were harvested from the wells by scraping with a 1mL pipette tip, washed, and stained with DAPI (1:100). The live cells (DAPI-negative) were sorted by the Westmead Institute for Medical Research (WIMR) using the BD FACSAria^TM^ III Cell Sorter and subsequently loaded into HIVE^TM^ devices, following the manufacturer’s instructions [22] (Honeycomb Biotechnologies, Inc). 12 HIVE^TM^ devices were loaded with approximately 30,000 cells from each sample (individual butyrate-treated and untreated samples from 4 patients and 2 controls) in 1 mL of DPBS + 1% FBS followed by 3 mL of cell media (DPBS + 1% FBS).

HIVEs^TM^ were then placed onto a spin plate and spun at 30 x g for 3 minutes to allow the single-cells to settle into picowells containing 3’ transcript-capture beads. The solution was removed from the HIVE^TM^ devices, and 2mL sample wash solution (provided in kit) was added to each. The sample wash solution was removed, and the cell-loaded HIVEs™ were frozen at −80°C after the addition of 2mL cell preservation solution (provided in kit), before being transferred to Australian Genome Research Facility (AGRF Ltd, Westmead) and processed through to single-cell NGS libraries [22].

Following standard protocol, cell-loaded HIVE^TM^ devices were sealed with a semi-permeable membrane, allowing for the use of the strong lysis solution followed by the addition of hybridization solution. After collection, beads with captured transcripts were extracted from the HIVE^TM^ device by centrifugation. The remaining HIVE^TM^ library preparation steps were conducted in a 96-well plate format [23]. The size distribution and quality of the final libraries were determined on a TapeStation 2000 platform with HD5000 ScreenTape System (Agilent Technologies, Santa Clara, CA, USA). The concentration of final pooled libraries was determined by qPCR. HIVE^TM^ scRNAseq libraries were sequenced using specific primers contained in the kit on an Illumina® NovaSeq® X platform (AGRF Ltd, Melbourne).

### Single-cell RNA sequencing bioinformatic analysis

Normalisation was performed using SCTransform and immune cell types were assigned with scType and scPred. We focused on broad immune cell types including T cells, B cells monocytes, eosinophils, myeloid dendritic cells and natural killer cells. Interestingly, scType results suggest that butyrate has a unique effect on CD4 T cell transcriptomes, suppressing expression of immune genes often used in scRNA-seq to identify CD4 T cells. We therefore clustered CD4 and CD8 T cells together as one cell population, rather than separating the populations for analysis. Merged data were then split by cell type and separately normalised, scaled, and integrated between patients using *harmony*, then UMAP (uniform manifold approximation and *projection)* projections were made using the first 30 dimensions. Differentially expressed genes were identified using *FindMarkers*.

### ORA pathways analysis and data presentation

Pathway enrichment analysis was performed via Over Representation Analysis (ORA) using the *clusterProfiler* package. DEGs were tested for pathway enrichment (FDR < 0.05) using Gene Ontology (GO) terms. Bar and dot plots of ORA GO results were plotted using *ggplot2* package. The top 5 up-regulated and down-regulated ORA GO post-simplified pathways for all cell types (bulk) were selected as representative GO terms. The top 10 up-regulated and down-regulated ORA GO pathways for T cells and bulk cells are presented on the dot plots. We analysed comparisons with all cells (bulk) as well as with the T cell population only. T cells were the largest individual cell population across our samples, representing 52-83% of the cell population per sample. Other cell populations were too small to analyse separately for statistically significant results.

To further evaluate themes within pathways, gene subclusters were created based on the most significant GO pathways per gene set and connectivity network (CNET) plots were created using the *enrichplot* package. The enriched pathways were represented by their respective colours and corresponding genes associated with the pathway.

To investigate the system-wide effects of butyrate, functional network grouping was conducted using a similarity matrix generated through the *pairwise_termsim* function and visualised with the *emapplot* function from the *enrichplot* package. The top 10 enriched GO terms from each ontology (Biological Process, Molecular Function, and Cellular Component) were organised into a network, with edges representing the overlap between gene sets, and plotted onto an enrichment map.

### Ethics Approval

Ethical approval was granted by the Sydney Children’s Hospitals Network Human Research Ethics Committee (HREC/18/SCHN/227, 2021/ETH00356). All families provided written informed consent for the study.

## Results

### Single-Cell RNA Sequencing (scRNA-seq): Differentially expressed genes

A total of 101,539 cells were sequenced across 10 samples from four patients with different NDDs at baseline (untreated, media only) and after *in vitro* butyrate treatment (media + 5mM butyrate), and two matched healthy controls (untreated, media only). Uniform manifold approximation and projection (UMAP) analysis of biological samples revealed 6 distinct cell clusters (Supplementary Figure 1). We compared ‘untreated patient’ versus ‘untreated control’ to determine the ‘baseline’ abnormality, and ‘butyrate-treated patient’ versus ‘untreated patient’, to determine the effect of butyrate on patient cells. These comparisons yielded 136 to 6,909 DEGs per comparison in the bulk cell analysis, and 175 to 2,229 DEGs per comparison in the T cell analysis (Supplementary Table 1).

### scRNA-seq in case vs control PBMCs

We compared the gene expression of each individual patient at baseline (untreated) against control (untreated) and performed pathway analysis (ORA GO), for all cell types combined (bulk) and T cells only. Our primary focus was on pathway analysis in the bulk (presented in main text) and T cells (presented in Supplementary Figure 2).

We found common themes of ribosomal, rRNA, and immune pathway dysregulation in the three monogenic NDD and the non-monogenic NDD patients, compared to healthy controls (Figure 1 and 2).

**Fig. 1.**
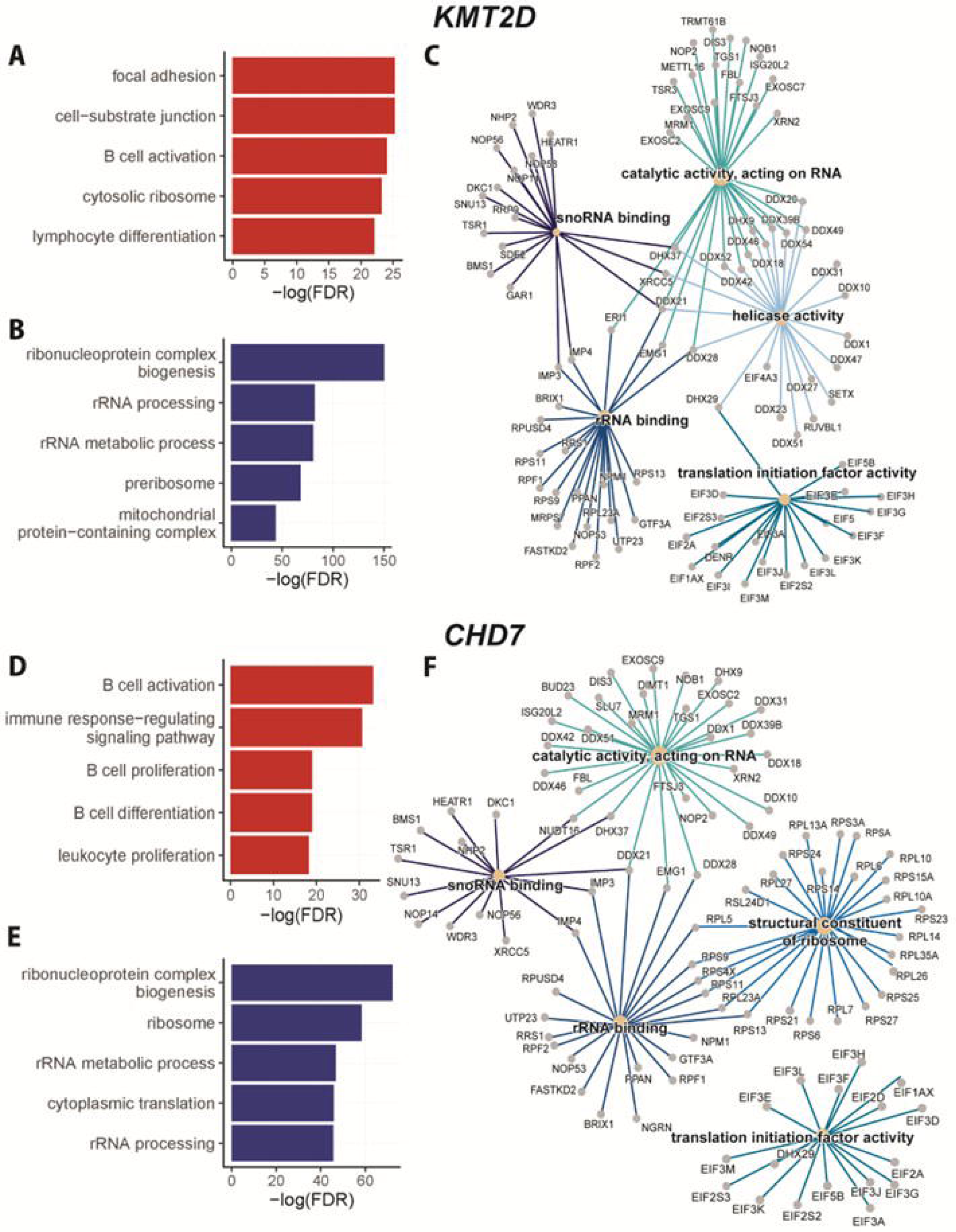
Single-cell RNA sequencing of PBMCs from children with NDD associated with monogenic variants in *KMT2D* and *CHD7*: bulk cell analysis of top 5 ORA GO pathways. The comparison of *KMT2D* versus control is presented as bar plots for **(A)** the top 5 upregulated ORA GO pathways (red), including adhesion, immune and ribosomal pathways, and **(B)** the top 5 downregulated ORA GO pathways (blue) including ribosomal and mitochondrial pathways. **(C)** A connectivity network (CNET) enrichment plot presents the top downregulated pathway for *KMT2D*, ‘ribonucleoprotein complex biogenesis’. Subcluster analysis via GO molecular function revealed themes of ‘catalytic activity, acting on RNA’ (enriched by *EXOSC* genes), ‘snoRNA binding’ (enriched by *NOP* genes), ‘rRNA binding’ (enriched by *RP* genes), ‘translation initiation factor activity’ (enriched by *EIF* genes) and ‘helicase activity’ (enriched by *DDX* genes). The comparison of *CHD7* versus control is presented as bar plots for **(D)** the top 5 upregulated ORA GO pathways (red), including immune pathways, and **(E)** top 5 downregulated ORA GO pathways (blue) including ribosomal and rRNA pathways. Similarly to *KMT2D*, **(F)** presents a CNET enrichment plot of the top downregulated pathway for *CHD7*, ‘ribonucleoprotein complex biogenesis’. Again, subcluster analysis via GO molecular function revealed themes of ‘catalytic activity, acting on RNA’ (enriched by *EXOSC* and *DDX* genes), ‘snoRNA binding’ (enriched by *NOP* genes), ‘rRNA binding’ (enriched by *RPF* genes), ‘translation initiation factor activity’ (enriched by *EIF* genes) and ‘structural constituent of ribosome’ (enriched by *RPS* and *RPL* genes).

**Fig. 2.**
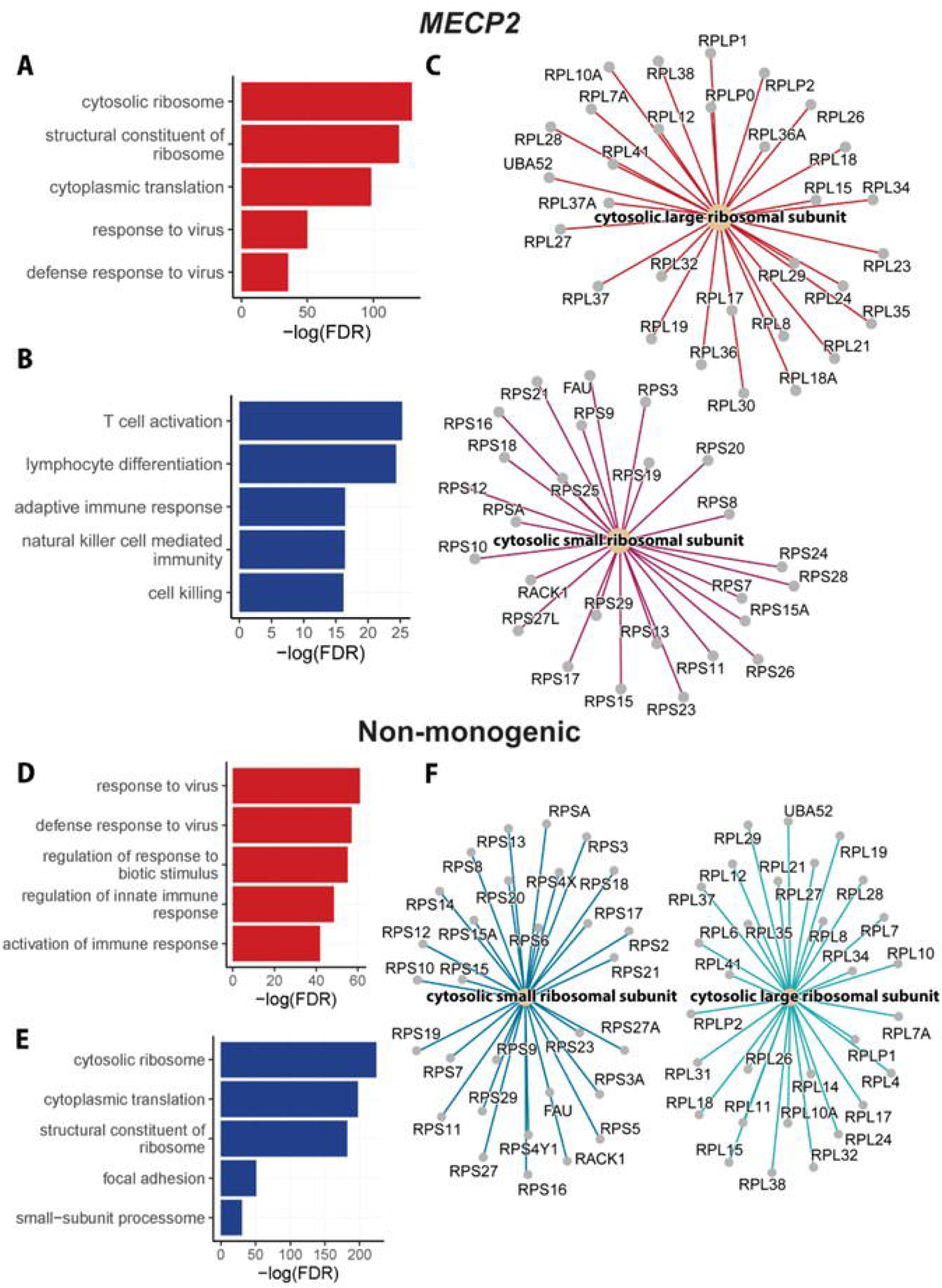
Single-cell RNA sequencing of PBMCs from a child with NDD associated with pathogenic variant in MECP2, and a child with no definable monogenic variant (non-monogenic): bulk cell analysis of top 5 GO pathways. The comparison of *MECP2* versus control is presented as bar plots for **(A)** the top 5 upregulated ORA GO pathways (red), including immune and ribosomal pathways, and **(B)** the top 5 downregulated ORA GO pathways (blue) including immune pathways. **(C)** A connectivity network (CNET) enrichment plot presents the top upregulated GO pathway for *MECP2*, ‘cytosolic ribosome’. Subcluster analysis via GO cellular component revealed themes of small ribosomal subunit (enriched by *RPS* genes) and large ribosomal subunit (enriched by *RPL* genes). The comparison of non-monogenic and control are presented as bar plots for **(D)** the top 5 upregulated ORA GO pathways (red), including immune pathways, and **(E)** the top 5 downregulated ORA GO pathways (blue), including ribosomal pathways. **(F)** A CNET enrichment plot presents the top downregulated pathways for the non-monogenic patient v control, ‘cytosolic ribosome’. Subcluster analysis via GO cellular component revealed themes of small ribosomal subunit (enriched by *RPS* genes) and large ribosomal subunit (enriched by *RPL* genes).

#### KMT2D (Kabuki syndrome)

*KMT2D* encodes a lysine (histone) methyltransferase called *MLL2* which is required for di- and tri-methylation of histone 3 lysine 4, central to histone methylation and chromatin arrangement [24]. Thus, pathological variants in *KMT2D* disrupt this mechanism, promoting a closed chromatin structure and limiting gene transcription [25]. Comparing PBMCs from the patient with Kabuki syndrome (*KMT2D)* vs control, the top five upregulated GO pathways included adhesion, immune and ribosomal pathways (Figure 1A). The top five downregulated GO pathways included ribosomal and mitochondrial pathways (Figure 1B). We focused on the most downregulated pathway in KMT2D vs control, which was the ‘ribonucleoprotein complex biogenesis’ pathway. Genes in this pathway (Figure 1C) revealed GO molecular function subclusters including ‘catalytic activity, acting on RNA’ (enriched by *EXOSC* genes), ‘snoRNA binding’ (enriched by *NOP* ribonucleoprotein genes), ‘rRNA binding’ (enriched by ribosomal protein *RP* genes), ‘translation initiation factor activity’ (enriched by *EIF* genes) and ‘helicase activity’ (enriched by *DDX* genes).

#### CHD7 (CHARGE syndrome)

The *CHD7* gene encodes the chromodomain helicase DNA binding protein 7, which is involved in regulating gene expression through chromatin remodelling [26]. Comparing PBMCs from a patient with CHARGE syndrome (*CHD7)* vs control, the top five upregulated GO pathways were all immune pathways (Figure 1D). The top five downregulated GO pathways included ribosomal and rRNA pathways (Figure 1E). We focused on the most downregulated pathway in *CHD7* vs control, which was also the ‘ribonucleoprotein complex biogenesis’ pathway, the same as for *KMT2D*. Genes in this *CHD7* ‘ribonucleoprotein complex biogenesis’ pathway (Figure 1F) revealed GO molecular function subclusters similar to *KMT2D*, including ‘catalytic activity, acting on RNA’ (enriched by *EXOSC* and *DDX* genes), ‘snoRNA binding’ (enriched by *NOP* genes), ‘rRNA binding’ (enriched by *RPF* genes), ‘translation initiation factor activity’ (enriched by *EIF* genes) and ‘structural constituent of ribosome’ (enriched by short and long ribosomal protein *RP* genes).

#### MECP2 (Rett syndrome)

The *MECP2* gene encodes the methyl-CpG binding protein 2, which is a transcription factor and chromatin-associated protein [27,28]. Comparing PBMCs from a patient with the NDD Rett syndrome (*MECP2)* vs control, the top five upregulated GO pathways were primarily ribosomal and immune pathways (Figure 2A). The top five downregulated GO pathways were immune pathways (Figure 2B). We focused on the genes from the upregulated ‘cytosolic ribosome pathway’ (Figure 2C), which revealed GO cellular component subclusters including cytosolic large ribosomal subunit (enriched by *RPL* genes) and cytosolic small ribosomal subunit (enriched by *RPS* genes).

### Non-monogenic NDD

The fourth patient had a complex NDD including autistic regression, attention deficit hyperactivity disorder, obsessive-compulsive disorder and Tourette syndrome. Exome sequencing had failed to identify a pathogenic variant (Table 1). Comparing PBMCs from this non-monogenic patient vs control, the top five upregulated GO pathways were immune pathways (Figure 2D). The top downregulated pathways were predominantly ribosomal pathways (Figure 2E). We focused on the downregulated ‘cytosolic ribosome pathway’ (Figure 2F), which revealed GO cellular component subclusters including cytosolic large ribosomal subunit (enriched by *RPL* genes) and cytosolic small ribosomal subunit (enriched by *RPS* genes).

In summary, at baseline (media only), there were common themes of immune, RNA and ribosomal dysregulation in all four NDD patients, including the non-monogenic patient.

### Effects of in vitro butyrate treatment on PBMCs derived from children with NDDs

We present the similarities and differences across *KMT2D* (Figure 3A), *CHD7* (Figure 3B), *MECP2* (Figure 3C), and non-monogenic NDD (Figure 3D) at baseline (case vs control) and after butyrate treatment (case-butyrate vs case) (Figures 3A-D). In general, there was similar enrichment of GO pathways between bulk and T cell comparisons across all four patient samples. Exploring the effects of butyrate, the baseline upregulated immune pathways in *KMT2D* (Figure 3A)*, CHD7* (Figure 3B) and non-monogenic (Figure 3D) patients were downregulated after butyrate treatment. The baseline downregulated ribosomal and rRNA processing pathways in *KMT2D* (Figure 3A)*, CHD7* (Figure 3B) and non-monogenic (Figure 3D) cells were upregulated after butyrate treatment.

**Fig. 3.**
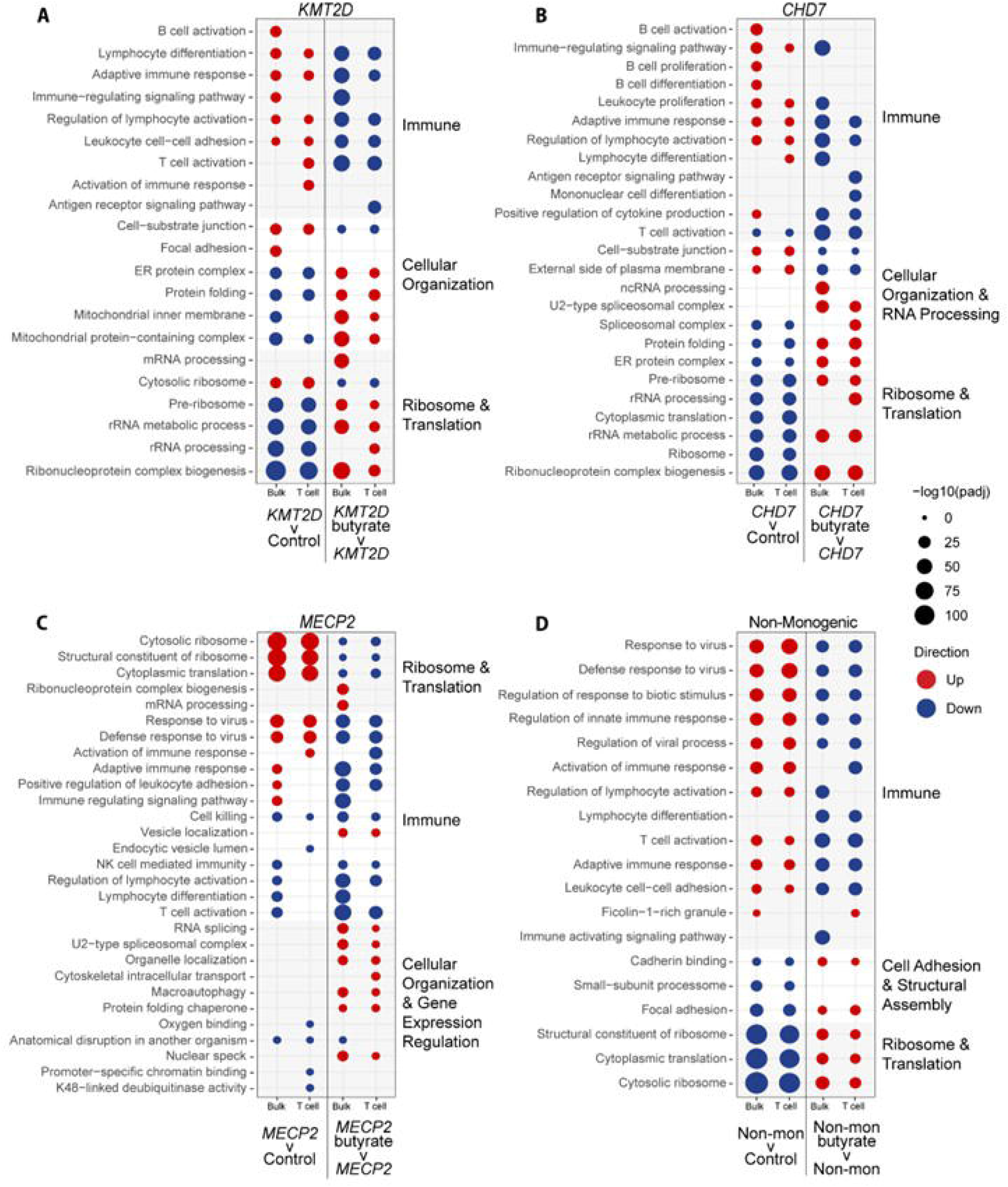
Dot plots visualising the top 10 upregulated and downregulated ORA GO pathways in untreated patient versus control PBMCs, and butyrate-treated patient cells versus untreated patient PBMCs. Both bulk analysis and T cell analysis are included, as labelled on the x-axis. The left-hand columns present the untreated patient versus control pathways, and the right-hand columns present the butyrate-treated versus untreated patient pathways. Red dots represent upregulated pathways, and blue dots represent downregulated pathways. The size of the dot represents the significance of pathway enrichment determined by -log10(padj) value, where padj is the adjusted p value. The pathways are clustered together based on similar function and then ranked according to enrichment. The pathway clusters are indicated by shading on the dot plot and labels to the right of the dot plot. There are separate plots for each NDD patient: (A) *KTM2D*, (B) *CHD7*, (C) *MECP2*, (D) Non-monogenic.

By contrast, for *MECP2* (Figure 3C), there was less consistent modification of pathways, although the baseline upregulated ribosomal pathways and dysregulated immune pathways were generally downregulated by butyrate.

### Systems-wide effects of butyrate treatment

Next, we explored system-wide effects of butyrate treatment on the non-monogenic PBMCs (bulk analysis) (Figure 4A). Butyrate treatment (non-monogenic butyrate vs non-monogenic) upregulated GO pathways relating to ribosome/translation, GTPase function, cytoskeleton, and mitochondrial function (in red), and downregulated epigenetic (histone, transcription activity) and immune GO pathways (in blue). Furthermore, we found that butyrate treatment normalised immune-related gene expression of important selected immune genes *IFITM3, CX3CR1, ISG15 and FCGR3A* in patient cells (Figure 4B).

**Fig. 4.**
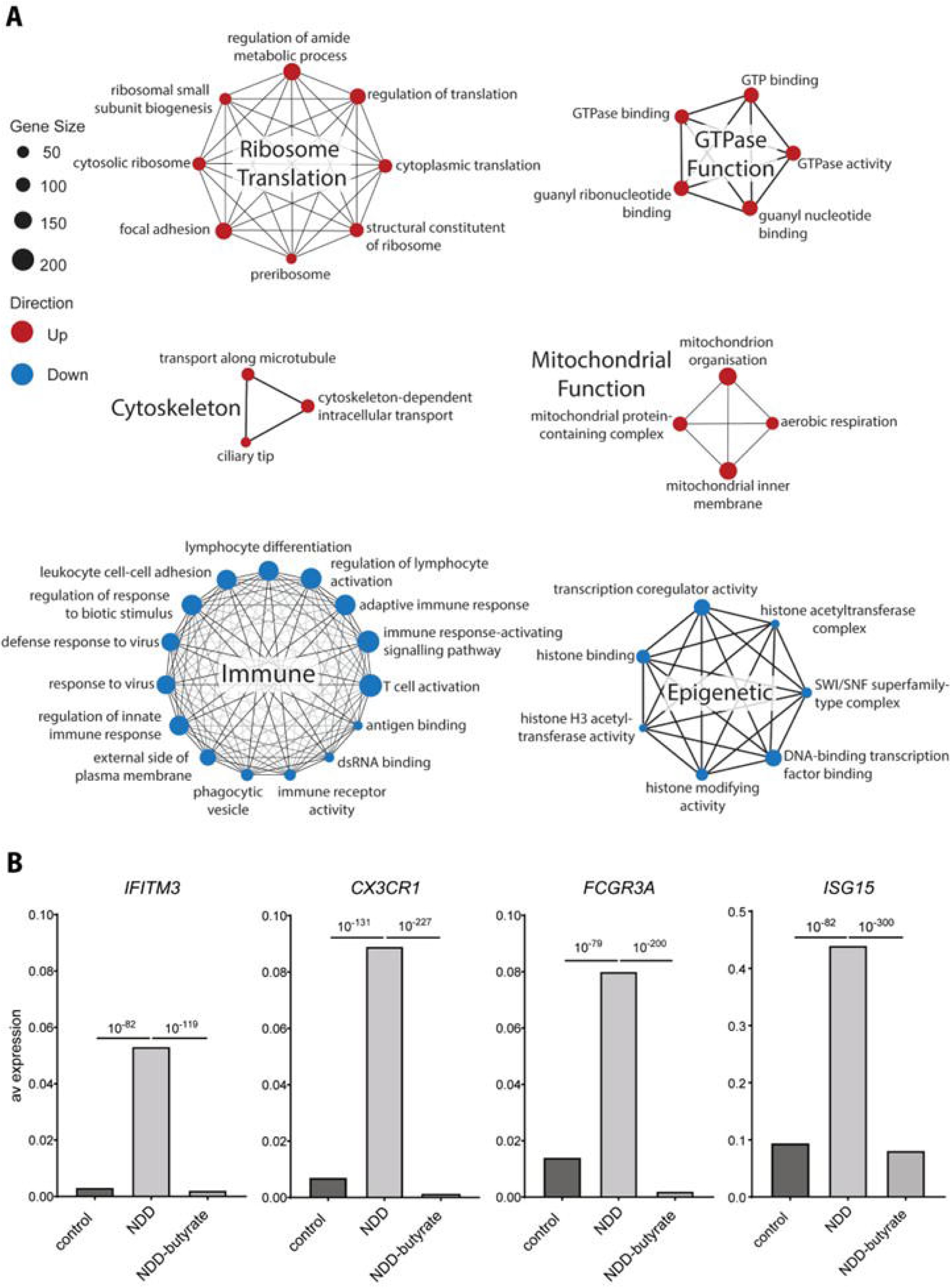
System wide effects of butyrate treatment on PBMCs from the child with non-monogenic NDD. **(A)** Functional network groupings of top 10 enriched GO terms from each ontology (biological process, molecular function and cellular component) in butyrate-treated versus untreated PBMCs from the non-monogenic patient comparison. Gene set size (number of genes) is represented by the size of the dot and edges represent the overlap between gene sets. Upregulated GO pathways are represented in red and downregulated GO pathways are represented in blue. **(B)** Comparison of average expression of *IFITM3, CX3CR1, FCGR3A, ISG15* immune genes between control PBMCs, NDD patient PBMCs and butyrate-treated NDD patient PBMCs. Statistical significance (p value) is indicated as line above graphs. Note difference in y-axis for *ISG15*.

## Discussion

Epigenetic mechanisms are increasingly recognised as key contributors to NDDs, arising from *de novo* pathogenic DNA variations in chromatin-related genes or through the interplay of common gene variants and environmental factors [7,8,11,19,29]. We selected children with *de novo* mutations in key chromatin-related genes *KMT2D* (Kabuki syndrome)*, CHD7* (CHARGE syndrome) and *MECP2* (Rett syndrome) as models of chromatin dysregulation, as well as one child with non-monogenic NDD. Through scRNA-seq analysis, patient PBMCs showed dysregulation of ribosomal and immune pathways compared to controls, at baseline. Butyrate, a short-chain fatty acid and HDAC-inhibitor with anti-inflammatory properties [30,31], reversed the abnormalities observed at baseline. Pathway analysis revealed butyrate-induced upregulation of ribosome/translation, GTPase activity, cytoskeletal organization, mitochondrial pathways, and downregulation of epigenetic (histone, transcription activity) and immune pathways.

We observed that *KMT2D, CHD7*, and non-monogenic NDD cells showed similar transcriptional changes at baseline, characterised by downregulated ribosomal and upregulated immune pathways. Whilst Kabuki and CHARGE syndrome are clinically distinct, there is significant phenotypic overlap [26,32,33]. *CHD7* and *MLL2* (encoded by *KMT2D*) interact with the same transcriptional machinery [26], suggesting shared regulatory pathways that may account for their similar transcriptomic profiles seen in our study. Additionally, dysregulation of ribosomal and immune pathways in non-monogenic NDD cells is a common RNA seq signature that we and others have observed in previous NDD cohorts [7,8,29,34,35]. In contrast, *MECP2* cells showed more variable pathway regulation, possibly due to the multifunctional nature of MeCP2 as both a repressor and promoter of gene transcription [28]. We hypothesise that chromatin dysregulation is an important mechanism across NDDs and is associated with an RNA seq signature characterised by ribosomal and immune dysregulation.

Ribosomal gene expression and biogenesis are tightly regulated by epigenetic mechanisms such as histone modifications, linking chromatin state to the cell’s capacity for protein synthesis [36]. *CHD7* is a positive regulator of ribosomal RNA (rRNA) biogenesis. Depleting cells of CHD7 results in hypermethylation of the rDNA promoter, and reduction of 45S pre-rRNA levels, cell proliferation, and protein synthesis [37]. As a histone modifier, *KMT2D* regulates ribosomal gene transcription and translation, and its depletion or mutation disrupts ribosomal protein biogenesis [11,38]. Ribosomes are the site of mRNA translation and play a critical role in brain development and disruptions to their translational machinery are associated with NDDs [39]. Perturbations in ribosomal biogenesis can impair protein synthesis essential for dendritic growth and synaptic function, leading to altered neuronal connectivity characteristic of NDDs [39]. Environmental exposures such as infections and oxidative stress, can further disrupt ribosome production and function, compromising cellular homeostasis [40–42]. In our study, we observed significant dysregulation of *RPL* and *RPS* genes, which encode components of the 40S and 60S ribosomal subunits, across the patient samples. We also identified dysregulation in ribonucleoprotein genes which play important roles in translation initiation (*EIF* genes) [43], ribosome biogenesis (*RPF* and *DDX* genes) [44,45], RNA metabolism and stability (*DDX* genes) [45], and RNA degradation (*EXOSC* genes) [46].

Due to the central role of chromatin regulation in determining cell fate, Kabuki syndrome, CHARGE syndrome, and Rett syndrome share common features of multiorgan involvement, in addition to neurodisability [27,47]. KMT2D, CHD7 and MeCP2 expression in immune cells also predisposes affected children to recurrent infections and infection-triggered neurodevelopmental or neuropsychiatric regression [19]. In Kabuki syndrome, there is increased susceptibility to infection in up to 70% of patients, due to B cell dysfunction, hypogammaglobulinemia, and CD4+ T cell deficiency [48,49]. In CHARGE syndrome due to *CHD7* mutations, immune deficiency is variable but includes T cell lymphopenia, B cell dysfunction, hypogammaglobulinemia, or rare severe combined immunodeficiencies [49,50]. Rett syndrome, caused by *MECP2* mutations, is linked to elevated proinflammatory cytokines, altered microglia and macrophage activation [51,52]. Together, these findings highlight the role of chromatin-related gene dysfunction in driving both neurodevelopmental and immune abnormalities, emphasising the importance of epigenetic regulation in maintaining neuroimmune homeostasis in NDDs.

In this study, butyrate treatment reversed immune and ribosomal pathway dysregulation in *KMT2D, CHD7,* and non-monogenic patient cells, with more limited effects in *MECP2*. Butyrate exerts both systemic and central effects, making it a key modulator of neuroimmune and neuro-epigenetic processes. Butyrate suppresses inflammation by promoting regulatory T cells, inhibiting pro-inflammatory T cells and macrophages, and inducing tolerogenic dendritic cells through HDAC inhibition and downstream signalling [53,54]. Through its influence on the gut–brain axis, butyrate modulates brain function via vagal nerve activation, endocrine signalling, and immune pathways [55]. Butyrate crosses the blood-brain barrier and inhibits HDACs, increasing histone acetylation, chromatin accessibility, and neuroprotective gene expression [56]. These effects promote hippocampal neurogenesis, synaptic plasticity, and memory formation [56]. Butyrate also modulates glial cell function, by promoting an anti-inflammatory phenotype, suppressing NF-κB–driven pro-inflammatory cytokines, supporting the resolution of neuroinflammation [57].

Given that *de novo* pathogenic DNA variations in chromatin-related genes account for only a minority of NDDs, it was clinically important to investigate how butyrate could be applied more broadly to non-monogenic NDDs. We observed that butyrate upregulated the expression of ribosome translation, GTPase function, cytoskeleton and mitochondrial function, and downregulated immune and epigenetic pathways in the cells of the non-monogenic NDD patient. GTPase activity and cytoskeleton organization pathways are important mechanisms of chromatin remodelling [58]. GTPases play critical roles in regulating chromatin structure, and transcription by controlling the localisation of chromatin modifiers and actin dynamics [59,60,61]. Cytoskeletal organisation further influences chromatin accessibility by altering nuclear shape and tension [62]. The observed downregulation of epigenetic pathways, including histone binding and histone modifying activity, was likely due to butyrate’s previously described HDAC inhibitory function [53,54,56]. Based on our findings, butyrate demonstrates promising epigenetic, anti-inflammatory and immunomodulatory effects, supporting its potential as a therapeutic intervention for NDDs.

Clinically, the ketogenic diet represents the most established analogue of butyrate, as it generates the ketone body β-hydroxybutyrate [63]. However, adherence to the ketogenic diet is challenging due to the need for strict dietary compliance and medical monitoring of ketones [64]. Exogenous ketone supplementation has emerged as a more practical alternative, with ketone salts (e.g., sodium, potassium, and calcium butyrate), esters (e.g., D-β-hydroxybutyrate isoform), and monoesters ((*R)*-3-hydroxybutyl *(R)*-3-hydroxybutyrate) now commercially available [65,66]. These compounds can acutely elevate circulating β-hydroxybutyrate levels without requiring prolonged fasting or carbohydrate restriction [66]. Nonetheless, they are limited by poor bioavailability, first-pass hepatic metabolism, and a short half-life, necessitating multi-dosing [56]. Importantly, most evidence for their efficacy is derived from acute, single-dose studies conducted under controlled laboratory conditions [65,67], and the long-term, clinically relevant effects of sustained exogenous ketone use remain underexplored [68]. To address the limitations of conventional supplementation, various modified butyrate derivates have been developed to enhance bioavailability. These include esterified or starch-conjugated forms such as serine-butyrate [69,70], high-amylose maize-resistant starch modified with acetate and butyrate (HAMSAB) [71], and N-(1-carbamoyl-2-phenyl-ethyl) butyramide (FBA) [72]. Further studies are required to evaluate the therapeutic efficacy, optimal dosing, and HDAC-inhibitory potential of butyrate and its derivates in human NDDs.

Limitations of this study include the small sample size and the use of scRNA-seq on PBMCs rather than brain tissue. However, due to the ubiquitous nature of chromatin in most cells, RNA sequencing of PBMCs make valuable and feasible potential biomarkers [11,35,73,74]. scRNA-seq enables high-resolution transcriptomic profiling of PBMCs, capturing cell-type-specific and treatment-related differences with strong statistical power. Another limitation of our study is that we focused on bulk and the larger cell population (T cells) for our analysis. The other cell populations (B cells, monocytes, natural killer cells, etc.) were too small to yield statistically powerful results across the comparison conditions. Additionally, targeted epigenetic, ATAC sequencing, histone modification profiling, and methylation analysis, are needed to explore the role of epigenetics in NDDs and the impact of butyrate. Also, detailed functional immune testing is required to confirm cellular immune deficits in NDDs, and their modification by butyrate. Future randomised clinical trials and *in vivo* studies, including different doses and duration, are needed to evaluate the safety, therapeutic efficacy, and biological impact of butyrate in improving symptoms of NDDs.

Overall, our study shows that children with NDDs, both with and without DNA variants in chromatin-related genes, share transcriptional abnormalities in peripheral immune cells. We propose that chromatin dysregulation leads to a common RNA seq signature involving ribosomal pathways with associated immune dysregulation. Chromatin dysregulation may represent a common mechanism across NDDs, affecting both brain and immune cells [7,8,11,29]. *In vitro* treatment with butyrate largely reversed these gene expression abnormalities, highlighting its potential as a therapeutic modulator of epigenetic and immune dysfunction in NDDs.

---

**Supplementary Table 1.**
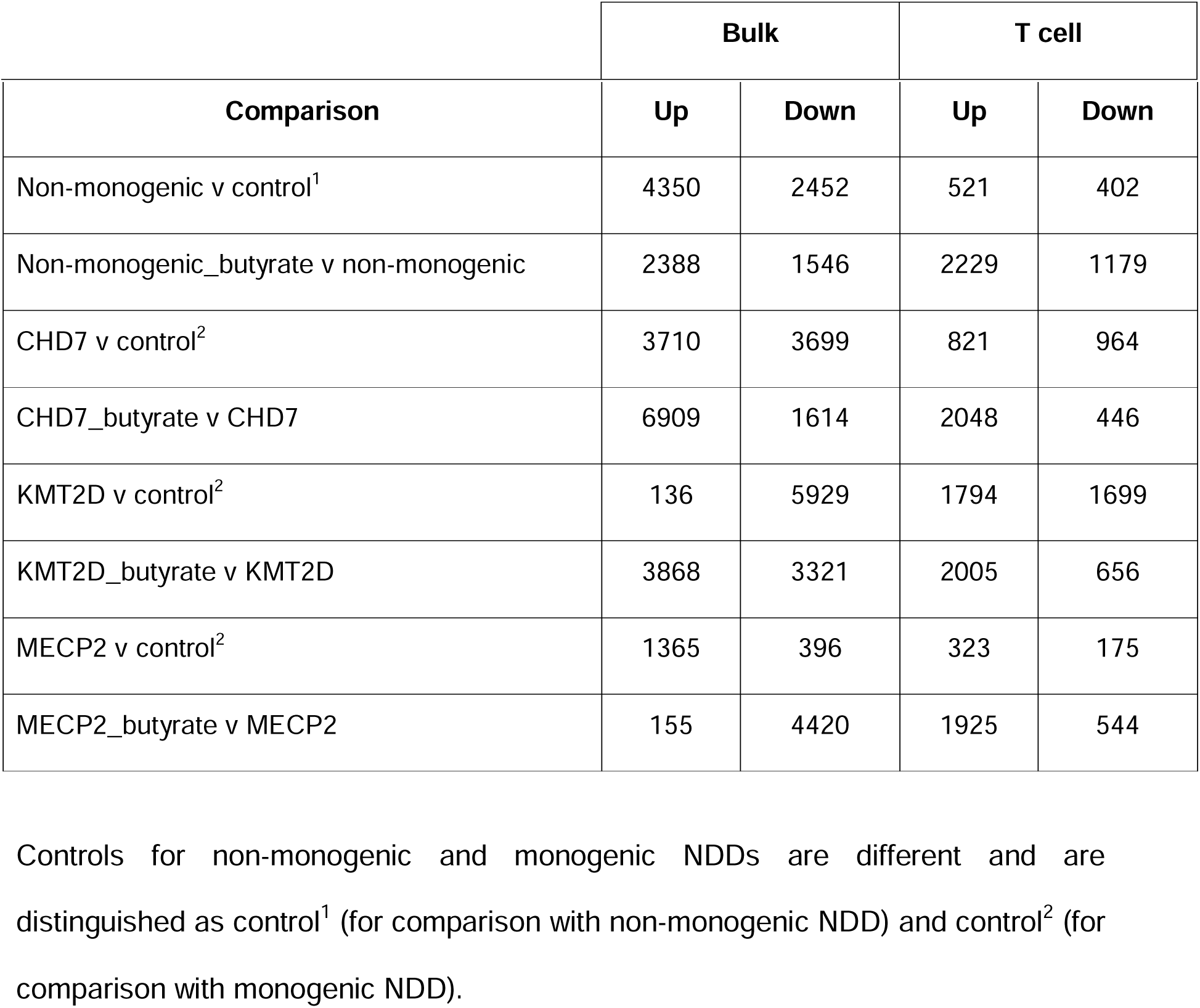
The number of up- and down-regulated differentially expressed genes (DEGs) of untreated and butyrate-treated PBMCs (bulk) and T cells from NDD patients and controls.

**Supplementary Fig. 1 UMAP and Quality Control Plots from scRNA-seq of NDD patient and control PBMCs. (A)** Uniform manifold approximation and projection (UMAP) analysis of 12 samples (including 4 children with monogenic and non-monogenic NDDs and 2 healthy controls) in scRNA-seq identified 6 distinct cell clusters including B cells (light blue), classical monocytes (grey), eosinophils (orange), myeloid dendritic cells (gold), natural killer cells (dark blue), and T cells (aqua), **(B)** Proportion of individual cell types across samples. **(C)** Heat map of the expression of identifying cell-surface markers for each cell cluster. Quality of samples is presented in the following violin plots: **(D)** Number of features, **(E)** Number of counts, and **(F**) mitochondrial percentage per sample.

**Supplementary Fig. 2 Single-cell RNA sequencing of PBMCs from children with NDDs: T cell analysis of top 5 ORA GO pathways.** The top 5 upregulated (red) and top 5 downregulated (blue) ORA GO pathways for each comparison are presented on bar plots. The significance of enrichment of the pathways is determined by -log(FDR). Note the different values on the x-axes. For *KMT2D* versus control, **(A)** presents the top upregulated pathways including adhesion, immune and ribosomal pathways, and **(B)** presents the top downregulated, including ribosomal pathways. For *CHD7* versus control **(C)** presents the top upregulated pathways, including immune pathways, and **(D)** presents the top downregulated pathways, including ribosomal and rRNA pathways. For *MECP2* versus control, **(E)** presents the top upregulated pathways, including immune and ribosomal pathways, and **(F)** and the top downregulated pathways including protein regulation, cytoskeletal, metabolic, gene regulation, and immune pathways. For non-monogenic versus control **(G)** presents the top upregulated pathways including immune pathways, and **(H)** presents the top downregulated pathways including ribosomal pathways.

## Resource availability

Anonymised data not published within this article will be made available by request from any qualified investigator. scRNA-seq data was uploaded to https://www.ncbi.nlm.nih.gov/geo/query/acc.cgi?acc=GSE296038 [access token: cnotcokedzuprcv].

## Data Availability

Anonymised data not published within this article will be made available by request from any qualified investigator. All published data are available online at https://www.ncbi.nlm.nih.gov/geo/query/acc.cgi?acc=GSE296038

## Acknowledgements

We would like to thank the patients and their families for participating in this study.

## Author contributions

HN, BK and RCD conceptualised and designed the experiments. JH, VXH, HN, ET, XL, RD, BK and SP performed the experiments and data acquisition. VXH and BG performed bioinformatic analysis. JH, VXH, HN, ET, MW, CE, BK, SM, MH, PV, SP, RCD analysed and interpreted the data. JH, VXH and RCD wrote the original manuscript. All authors critically revised and edited the manuscript. All authors approved the final version of the article.

## References

1. De Rubeis S, He X, Goldberg AP, Poultney CS, Samocha K, Ercument Cicek A, et al. Synaptic, transcriptional and chromatin genes disrupted in autism. Nature. 2014 Nov;515(7526):209–15.

2. Yan J, Kothur K, Mohammad S, Chung J, Patel S, Jones HF, et al. CSF neopterin, quinolinic acid and kynurenine/tryptophan ratio are biomarkers of active neuroinflammation. eBioMedicine. 2023 Apr 27;91:104589.

3. Tiffon C. The Impact of Nutrition and Environmental Epigenetics on Human Health and Disease. Int J Mol Sci. 2018 Nov 1;19(11):3425.

4. Salinas RD, Connolly DR, Song H. Epigenetics in Neurodevelopment. Neuropathol Appl Neurobiol. 2020 Feb;46(1):6–27.

5. Cavalli G, Heard E. Advances in epigenetics link genetics to the environment and disease. Nature. 2019 Jul;571(7766):489–99.

6. Cooper GM. Chromosomes and Chromatin. In: The Cell: A Molecular Approach 2nd edition [Internet]. Sinauer Associates; 2000 [cited 2025 Apr 4]. Available from: https://www.ncbi.nlm.nih.gov/books/NBK9863/

7. Nishida H, Han VX, Keating BA, Zyner KG, Gloss B, Aryamanesh N, et al. Chromatin, transcriptional and immune dysregulation in children with neurodevelopmental regression [Internet]. medRxiv; 2025 [cited 2025 Apr 3]. p. 2025.03.05.25322433. Available from: https://www.medrxiv.org/content/10.1101/2025.03.05.25322433v1

8. Han VX, Nishida H, Keating BA, Gloss BS, Lau X, Dissanayake R, et al. Intravenous immunoglobulin has epigenetic, ribosomal, and immune effects in Paediatric Acute-Onset Neuropsychiatric Syndrome [Internet]. medRxiv; 2025 [cited 2025 May 20]. p. 2025.03.27.25324808. Available from: https://www.medrxiv.org/content/10.1101/2025.03.27.25324808v1

9. Park SY, Kim JS. A short guide to histone deacetylases including recent progress on class II enzymes. Exp Mol Med. 2020 Feb;52(2):204–12.

10. Berni Canani R, Di Costanzo M, Leone L. The epigenetic effects of butyrate: potential therapeutic implications for clinical practice. Clin Epigenet. 2012 Dec;4(1):1–7.

11. Tsang E, Han VX, Flutter C, Alshammery S, Keating BA, Williams T, et al. Ketogenic diet modifies ribosomal protein dysregulation in KMT2D Kabuki syndrome. eBioMedicine. 2024 May 19;104:105156.

12. Meltzer A, Van de Water J. The Role of the Immune System in Autism Spectrum Disorder. Neuropsychopharmacol. 2017 Jan;42(1):284–98.

13. Jiang NM, Cowan M, Moonah SN, Petri WA. The Impact of Systemic Inflammation on Neurodevelopment. Trends Mol Med. 2018 Sep;24(9):794–804.

14. Sreenivas N, Maes M, Padmanabha H, Dharmendra A, Chakkera P, Paul Choudhury S, et al. Comprehensive immunoprofiling of neurodevelopmental disorders suggests three distinct classes based on increased neurogenesis, Th-1 polarization or IL-1 signaling. Brain, Behavior, and Immunity. 2024 Jan 1;115:505–16.

15. Han VX, Jones HF, Patel S, Mohammad SS, Hofer MJ, Alshammery S, et al. Emerging evidence of Toll-like receptors as a putative pathway linking maternal inflammation and neurodevelopmental disorders in human offspring: A systematic review. Brain, Behavior, and Immunity. 2022 Jan 1;99:91–105.

16. Martino D, Dale RC, Gilbert DL, Giovannoni G, Leckman JF. Immunopathogenic mechanisms in tourette syndrome: A critical review. Movement Disorders. 2009;24(9):1267–79.

17. Gabriele M, Lopez Tobon A, D’Agostino G, Testa G. The chromatin basis of neurodevelopmental disorders: Rethinking dysfunction along the molecular and temporal axes. Progress in Neuro-Psychopharmacology and Biological Psychiatry. 2018 Jun 8;84:306–27.

18. Faundes V, Newman WG, Bernardini L, Canham N, Clayton-Smith J, Dallapiccola B, et al. Histone Lysine Methylases and Demethylases in the Landscape of Human Developmental Disorders. Am J Hum Genet. 2018 Jan 4;102(1):175–87.

19. Dale RC, Mohammad S, Han VX, Nishida H, Goel H, Tangye SG, et al. Pathogenic variants in chromatin-related genes: Linking immune dysregulation to neuroregression and acute neuropsychiatric disorders. Developmental Medicine & Child Neurology. 2025 Feb 22;00:1–8.

20. Thammongkol S, Vears DF, Bicknell-Royle J, Nation J, Draffin K, Stewart KG, et al. Efficacy of the ketogenic diet: Which epilepsies respond? Epilepsia. 2012;53(3):e55–9.

21. Qiao YN, Li L, Hu SH, Yang YX, Ma ZZ, Huang L, et al. Ketogenic diet-produced β-hydroxybutyric acid accumulates brain GABA and increases GABA/glutamate ratio to inhibit epilepsy. Cell Discov. 2024 Feb 13;10:17.

22. Honeycomb Biotechnologies. HIVE CLX scRNAseq Solution. [cited 2024 Oct 1]. HIVE CLX Sample Capture User Protocol. Available from: https://honeycombbio.zendesk.com/hc/en-us/articles/15173728631707-HIVE-CLX-Sample-Capture-User-Protocol

23. Honeycomb Biotechnologies. HIVE CLX scRNAseq Solution. [cited 2024 Oct 1]. HIVE CLX Transcriptome Recovery & Library Preparation User Protocol. Available from: https://honeycombbio.zendesk.com/hc/en-us/articles/15173667166491-HIVE-CLX-Transcriptome-Recovery-Library-Preparation-User-Protocol

24. Froimchuk E, Jang Y, Ge K. Histone H3 lysine 4 methyltransferase KMT2D. Gene. 2017 Sep 5;627:337–42.

25. Jung YL, Hung C, Choi J, Lee EA, Bodamer O. Characterizing the molecular impact of KMT2D variants on the epigenetic and transcriptional landscapes in Kabuki syndrome. Human Molecular Genetics. 2023 Jul 1;32(13):2251–61.

26. Schulz Y, Freese L, Mänz J, Zoll B, Völter C, Brockmann K, et al. CHARGE and Kabuki syndromes: a phenotypic and molecular link. Human Molecular Genetics. 2014 Aug 15;23(16):4396–405.

27. Amir RE, Van den Veyver IB, Wan M, Tran CQ, Francke U, Zoghbi HY. Rett syndrome is caused by mutations in X-linked MECP2, encoding methyl-CpG-binding protein 2. Nat Genet. 1999 Oct;23(2):185–8.

28. Vuu YM, Roberts CT, Rastegar M. MeCP2 Is an Epigenetic Factor That Links DNA Methylation with Brain Metabolism. International Journal of Molecular Sciences. 2023 Jan;24(4):4218.

29. Han VX, Alshammery S, Keating BA, Gloss BS, Hofer MJ, Graham ME, et al. Epigenetic, ribosomal, and immune dysregulation in Paediatric Acute-Onset Neuropsychiatric Syndrome [Internet]. medRxiv; 2025 [cited 2025 Apr 15]. p. 2025.03.28.25324649. Available from: https://www.medrxiv.org/content/10.1101/2025.03.28.25324649v1

30. Kratsman N, Getselter D, Elliott E. Sodium butyrate attenuates social behavior deficits and modifies the transcription of inhibitory/excitatory genes in the frontal cortex of an autism model. Neuropharmacology. 2016 Mar;102:136–45.

31. Takuma K, Hara Y, Kataoka S, Kawanai T, Maeda Y, Watanabe R, et al. Chronic treatment with valproic acid or sodium butyrate attenuates novel object recognition deficits and hippocampal dendritic spine loss in a mouse model of autism. Pharmacol Biochem Behav. 2014 Nov;126:43–9.

32. Butcher DT, Cytrynbaum C, Turinsky AL, Siu MT, Inbar-Feigenberg M, Mendoza-Londono R, et al. CHARGE and Kabuki Syndromes: Gene-Specific DNA Methylation Signatures Identify Epigenetic Mechanisms Linking These Clinically Overlapping Conditions. Am J Hum Genet. 2017 May 4;100(5):773–88.

33. Ufartes R, Grün R, Salinas G, Sitte M, Kahl F, Wong MTY, et al. CHARGE syndrome and related disorders: a mechanistic link. Human Molecular Genetics. 2021 Dec 1;30(23):2215–24.

34. Lombardo MV. Ribosomal protein genes in post-mortem cortical tissue and iPSC-derived neural progenitor cells are commonly upregulated in expression in autism. Mol Psychiatry. 2021 May;26(5):1432–5.

35. Gevezova M, Sbirkov Y, Sarafian V, Plaimas K, Suratanee A, Maes M. Autistic spectrum disorder (ASD) – Gene, molecular and pathway signatures linking systemic inflammation, mitochondrial dysfunction, transsynaptic signalling, and neurodevelopment. Brain Behav Immun Health. 2023 Jun 5;30:100646.

36. Denisenko O. Epigenetics of Ribosomal RNA Genes. Biochemistry Moscow. 2022 Jan 1;87(1):S103–10.

37. Zentner GE, Hurd EA, Schnetz MP, Handoko L, Wang C, Wang Z, et al. CHD7 functions in the nucleolus as a positive regulator of ribosomal RNA biogenesis. Hum Mol Genet. 2010 Sep 15;19(18):3491–501.

38. Xu J, Zhong A, Zhang S, Chen M, Zhang L, Hang X, et al. KMT2D Deficiency Promotes Myeloid Leukemias which Is Vulnerable to Ribosome Biogenesis Inhibition. Adv Sci (Weinh). 2023 May 4;10(19):2206098.

39. Hetman M, Slomnicki LP. Ribosomal biogenesis as an emerging target of neurodevelopmental pathologies. J Neurochem. 2019 Feb;148(3):325–47.

40. Shcherbik N, Pestov DG. The Impact of Oxidative Stress on Ribosomes: From Injury to Regulation. Cells. 2019 Nov 2;8(11):1379.

41. Ni C, Buszczak M. The homeostatic regulation of ribosome biogenesis. Semin Cell Dev Biol. 2023 Feb 28;136:13–26.

42. Wang X, Zhu J, Zhang D, Liu G. Ribosomal control in RNA virus-infected cells. Front Microbiol [Internet]. 2022 Nov 7 [cited 2025 May 16];13. Available from: https://www.frontiersin.org/journals/microbiology/articles/10.3389/fmicb.2022.1026887/ful l

43. Hao P, Yu J, Ward R, Liu Y, Hao Q, An S, et al. Eukaryotic translation initiation factors as promising targets in cancer therapy. Cell Communication and Signaling. 2020 Nov 4;18(1):175.

44. Dörner K, Ruggeri C, Zemp I, Kutay U. Ribosome biogenesis factors—from names to functions. EMBO J. 2023 Feb 10;42(7):e112699.

45. Abdelhaleem M, Maltais L, Wain H. The human *DDX* and *DHX* gene families of putative RNA helicases. Genomics. 2003 Jun 1;81(6):618–22.

46. Zhang W, Zhu J, He X, Liu X, Li J, Li W, et al. Exosome complex genes mediate RNA degradation and predict survival in mantle cell lymphoma. Oncol Lett. 2019 Nov;18(5):5119–28.

47. Bögershausen N, Gatinois V, Riehmer V, Kayserili H, Becker J, Thoenes M, et al. Mutation Update for Kabuki Syndrome Genes KMT2D and KDM6A and Further Delineation of X-Linked Kabuki Syndrome Subtype 2. Human Mutation. 2016;37(9):847– 64.

48. Lin JL, Lee WI, Huang JL, Chen PKT, Chan KC, Lo LJ, et al. Immunologic assessment and KMT2D mutation detection in Kabuki syndrome. Clinical Genetics. 2015;88(3):255– 60.

49. Hoffman JD, Ciprero KL, Sullivan KE, Kaplan PB, McDonald-McGinn DM, Zackai EH, et al. Immune abnormalities are a frequent manifestation of Kabuki syndrome. American Journal of Medical Genetics Part A. 2005;135A(3):278–81.

50. Liu ZZ, Wang ZL, Choi TI, Huang WT, Wang HT, Han YY, et al. *Chd7* Is Critical for Early T-Cell Development and Thymus Organogenesis in Zebrafish. The American Journal of Pathology. 2018 Apr 1;188(4):1043–58.

51. Jyonouchi S, McDonald-McGinn DM, Bale S, Zackai EH, Sullivan KE. CHARGE (Coloboma, Heart Defect, Atresia Choanae, Retarded Growth and Development, Genital Hypoplasia, Ear Anomalies/Deafness) Syndrome and Chromosome 22q11.2 Deletion Syndrome: A Comparison of Immunologic and Nonimmunologic Phenotypic Features. Pediatrics. 2009 May 1;123(5):e871–7.

52. Cronk JC, Derecki NC, Ji E, Xu Y, Lampano AE, Smirnov I, et al. Methyl-CpG binding protein 2 regulates microglia and macrophage gene expression in response to inflammatory stimuli. Immunity. 2015 Apr 21;42(4):679–91.

53. Kaisar MMM, Pelgrom LR, van der Ham AJ, Yazdanbakhsh M, Everts B. Butyrate Conditions Human Dendritic Cells to Prime Type 1 Regulatory T Cells via both Histone Deacetylase Inhibition and G Protein-Coupled Receptor 109A Signaling. Front Immunol. 2017 Oct 30;8:1429.

54. Sarkar A, Mitra P, Lahiri A, Das T, Sarkar J, Paul S, et al. Butyrate limits inflammatory macrophage niche in NASH. Cell Death Dis. 2023 May 18;14(5):332.

55. de Alpino G CÁ, Pereira-Sol Gabriela Amorim, Dias Mariana de Mourae, de Aguiar Aline Silva, and do Peluzio M CG. Beneficial effects of butyrate on brain functions: A view of epigenetic. Critical Reviews in Food Science and Nutrition. 2024 May 6;64(12):3961– 70.

56. Steliou K, Boosalis MS, Perrine SP, Sangerman J, Faller DV. Butyrate Histone Deacetylase Inhibitors. Biores Open Access. 2012 Aug;1(4):192–8.

57. Huuskonen J, Suuronen T, Nuutinen T, Kyrylenko S, Salminen A. Regulation of microglial inflammatory response by sodium butyrate and short-chain fatty acids. Br J Pharmacol. 2004 Mar;141(5):874–80.

58. Rajakylä EK, Vartiainen MK. Rho, nuclear actin, and actin-binding proteins in the regulation of transcription and gene expression. Small GTPases. 2014 Mar 6;5:e27539.

59. Dworak N, Makosa D, Chatterjee M, Jividen K, Yang C, Snow C, et al. A nuclear lamina-chromatin-Ran GTPase axis modulates nuclear import and DNA damage signaling. Aging Cell. 2019 Feb;18(1):e12851.

60. Farrants AKÖ. Chromatin remodelling and actin organisation. FEBS Letters. 2008 Jun 18;582(14):2041–50.

61. Moon SY, Zheng Y. Rho GTPase-activating proteins in cell regulation. Trends in Cell Biology. 2003 Jan 1;13(1):13–22.

62. Xie X, Mahmood SR, Gjorgjieva T, Percipalle P. Emerging roles of cytoskeletal proteins in regulating gene expression and genome organization during differentiation. Nucleus. 2020 Mar 25;11(1):53–65.

63. Dąbek A, Wojtala M, Pirola L, Balcerczyk A. Modulation of Cellular Biochemistry, Epigenetics and Metabolomics by Ketone Bodies. Implications of the Ketogenic Diet in the Physiology of the Organism and Pathological States. Nutrients. 2020 Mar 17;12(3):788.

64. Batch JT, Lamsal SP, Adkins M, Sultan S, Ramirez MN. Advantages and Disadvantages of the Ketogenic Diet: A Review Article. Cureus. 2020 Aug 10;12(8):e9639.

65. Falkenhain K, Daraei A, Forbes SC, Little JP. Effects of Exogenous Ketone Supplementation on Blood Glucose: A Systematic Review and Meta-analysis. Advances in Nutrition. 2022 Sep 1;13(5):1697–714.

66. Saris CGJ, Timmers S. Ketogenic diets and Ketone suplementation: A strategy for therapeutic intervention. Front Nutr. 2022 Nov 15;9:947567.

67. Stubbs BJ, Cox PJ, Evans RD, Santer P, Miller JJ, Faull OK, et al. On the Metabolism of Exogenous Ketones in Humans. Front Physiol. 2017 Oct 30;8:848.

68. Soto-Mota A, Vansant H, Evans RD, Clarke K. Safety and tolerability of sustained exogenous ketosis using ketone monoester drinks for 28 days in healthy adults. Regulatory Toxicology and Pharmacology. 2019 Dec 1;109:104506.

69. Cao S, Budina E, Raczy MM, Solanki A, Nguyen M, Beckman TN, et al. A serine-conjugated butyrate prodrug with high oral bioavailability suppresses autoimmune arthritis and neuroinflammation in mice. Nat Biomed Eng. 2024 May;8(5):611–27.

70. Beckman T, Volpatti L, Cao S, Slezak A, Reda J, Budina E, et al. A Pro-Drug Strategy Enhances Butyrate Bioavailability and Therapeutic Efficacy In Atherosclerosis. The Journal of Immunology. 2024 May 1;212(1_Supplement):0853_5126.

71. Bell KJ, Saad S, Tillett BJ, McGuire HM, Bordbar S, Yap YA, et al. Metabolite-based dietary supplementation in human type 1 diabetes is associated with microbiota and immune modulation. Microbiome. 2022 Jan 19;10:9.

72. Russo R, Santarcangelo C, Badolati N, Sommella E, De Filippis A, Dacrema M, et al. *In vivo* bioavailability and *in vitro* toxicological evaluation of the new butyric acid releaser N-(1-carbamoyl-2-phenyl-ethyl) butyramide. Biomedicine & Pharmacotherapy. 2021 May 1;137:111385.

73. Shen L, Feng C, Zhang K, Chen Y, Gao Y, Ke J, et al. Proteomics Study of Peripheral Blood Mononuclear Cells (PBMCs) in Autistic Children. Front Cell Neurosci [Internet]. 2019 Mar 19 [cited 2025 Apr 15];13. Available from: https://www.frontiersin.org https://www.frontiersin.org/journals/cellular-neuroscience/articles/10.3389/fncel.2019.00105/full

74. Goodman SJ, Luperchio TR, Ellegood J, Chater-Diehl E, Lerch JP, Bjornsson HT, et al. Peripheral blood DNA methylation and neuroanatomical responses to HDACi treatment that rescues neurological deficits in a Kabuki syndrome mouse model. Clinical Epigenetics. 2023 Oct 27;15(1):172.

