## Supplementary figures and images for "Butyrate modifies epigenetic and immune pathways in peripheral mononuclear cells from children with neurodevelopmental disorders associated with chromatin dysregulation"

### Supplementary figure 1

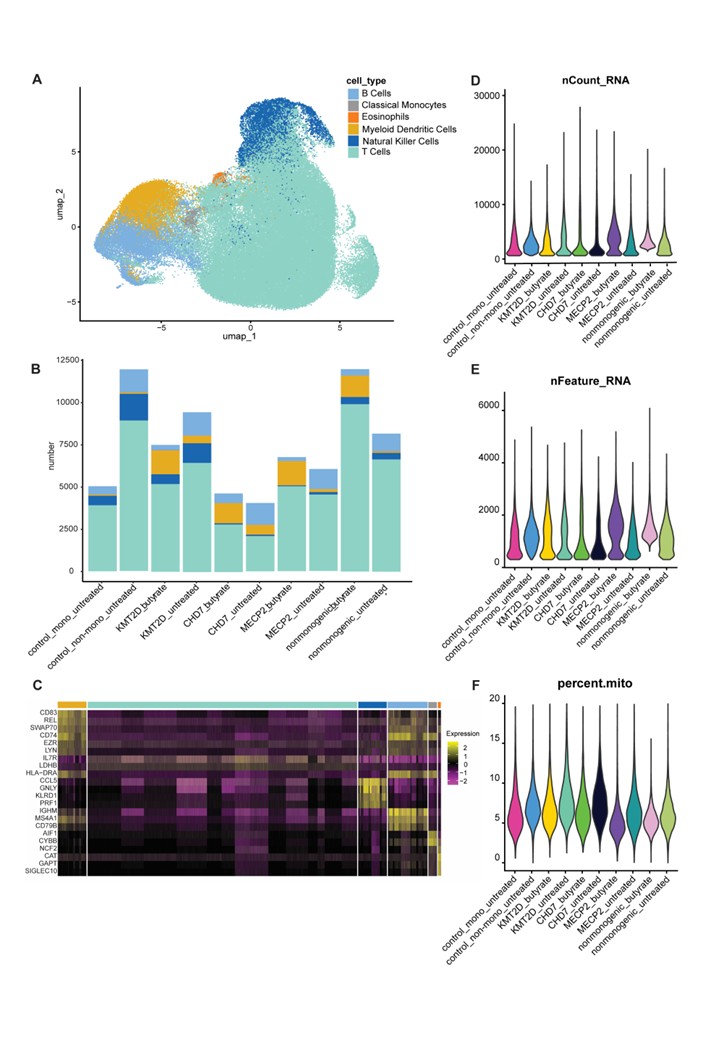

### Supplementary figure 2

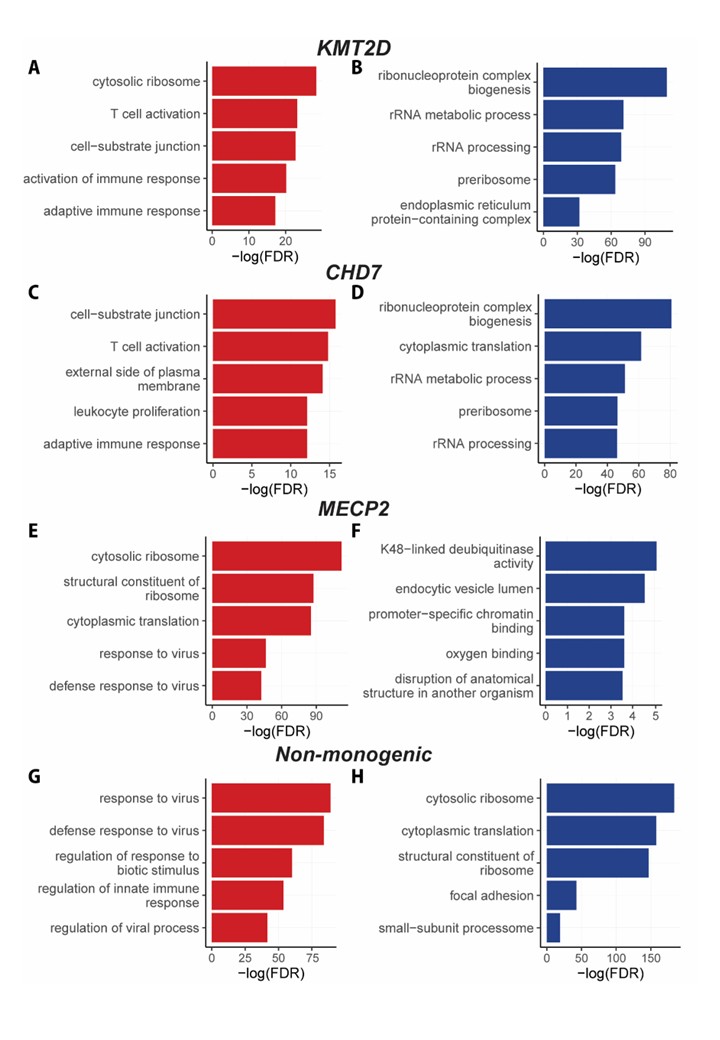
